# Testing for bi-directional rejection thresholds to sweetness and their links to sugar intake and sweet-taste drink consumption

**DOI:** 10.1101/2022.11.27.22282800

**Authors:** Mei Peng, Rachel Ginieis, Sashie Abeywickrema, Jessica McCormack, John Prescott

**Affiliations:** Sensory Neuroscience Laboratory, Department of Food Science, University of Otago, Dunedin, New Zealand; Department of Public Health and Basic Sciences, Faculty of Allied Health Sciences, University of Sri Jayewardenepura, Nugegoda, Sri Lanka; Department of Physiology, Faculty of Medicine, University of Colombo, Colombo 03, Sri Lanka; Taste Matters Research & Consulting, Australia; Department of Agriculture, Food, Environment and Forestry (DAGRI), University of Florence, Italy

**Author notes:** Corresponding author: Mei Peng, PhD, Department of Food Science, PO Box 56, Dunedin 9054, New Zealand.

**Keywords:** sweet taste, taste perception, sugar reduction, obesity

## Abstract

**Background and Aims:** Sugar intake has been linked to obesity, however, the relationship between individual sugar perception and dietary choice remains unclear. The current study aims to measure individual bi-directional rejection thresholds and compare these measures to sugar intake and consumption patterns of sweet-taste beverages.

**Methods:** A cross-section design will be used to analyse the relationship between sweetness perception, sucrose liking, and dietary intake. Participants will attend laboratory sessions to assess sucrose liking, ascending and descending rejection thresholds, and detection thresholds, and complete a 4-day weighed food-diary to assess dietary intake. ANCOVA will be used to test for differences in detection threshold and hedonic VAS ratings between the liking and disliking group, with age, gender, and BMI as covariates. A generalised linear mixed-models will be applied to test for differences in individual ascending versus descending rejection threshold across the two sweet liker status groups. Regression models will be used to test the role of ascending versus descending rejection thresholds on predicting sugar and sweet-taste beverage consumption.

**Discussion:** More research focusing on links between individual sweetness perception and sugar intake will be important for elucidating the mechanism underpinning sensory effects on dietary behaviour.

## 1. Introduction

In light of the current obesity epidemic, the link between sugar sweetened beverage consumption and obesity has been attracted substantial research attention (e.g., Pereira, 2014), with extensive results pointing to a positive relationship (Berkey, Rockett, Field, Gillman, & Colditz, 2004; Vasanti S. Malik & Hu, 2022; Vasanti S Malik, Pan, Willett, & Hu, 2013). These findings indicate a potential scope to investigate factors driving inter-individual differences in sugar consumption patterns.

It is widely accepted that humans like sweetness from birth, due to its association with the presence of energy (Ganchrow & Mennella, 2003). Nevertheless, people vary greatly in terms of their sweetness sensitivity (Peng, Hautus, Oey, & Silcock, 2016), intensity (Waksmonski & Koppel, 2016) and hedonic perception (Kim, Prescott, & Kim, 2014). Individuals can be grouped into sweet “likers” (SL) and “dislikers” (SD) (Garneau, Nuessle, Mendelsberg, Shepard, & Tucker, 2018; Kim et al., 2014; Kim, Prescott, & Kim, 2017), which sweet likers showing increased liking with increased sugar concentration, and dislikers decreased liking with increased sugar concentration..

The relationship between individual sweetness perception and dietary behaviour, in particularly sugar intake, is unclear. To date, findings to this question remain to be highly contrasting. While some studies observed strong taste-diet associations (Abeywickrema, Ginieis, Oey, & Peng, 2022; Jayasinghe et al., 2017), some others found a null relationship (Cicerale, Riddell, & Keast, 2012). To a large extent, these inconsistent results are attributable to different choices of psychophysical methods for quantifying individual sweet perception (Tan & Tucker, 2019). Relatively, hedonic or acceptance tests have shown more promise in terms of detecting a taste-diet association.

Rejection threshold (RjT) was proposed by Prescott, Norris, Kunst, and Kim (2005), initially as an approach to measure effects of taint (undesirable flavour note) on wine preference. This methodology had a rapid uptake due to its clear advantage in predicting product acceptance, especially when comparing to the classic measure of detection threshold. Methven, Xiao, Cai, and Prescott (2016) applied this methodology in a novel context, to estimate RjTs to a desirable taste, i.e., sweetness, across individuals. The study found little difference in RjTs across sweet likers versus dislikers, although the authors speculated that larger studies might produce differential results.

To date, RjT measures have only been used to assess effects of stimulus increments, which is termed *ascending* RjT in this paper. Little is known about effects of stimulus reductions on product liking. Building upon these contexts, the current study aims to test individual RjTs to both *ascending* and *descending* sucrose concentrations. These measures are compared against sweet liker status and dietary sugar intake.

## 2. Methods

### 2.1 Participants

A sample of 63 healthy participants will be recruited. A sample-size calculation was done based on effect sizes reported in previous studies testing for links between sweetness perception and sugar intake(Jayasinghe et al., 2017; Low, Lacy, & Keast, 2014). With 80% power and an α-level of 0.05, G*Power 3.1.9.7 Software estimated a sample size of 49 (effect size r = 0.35), plus fourteen participants in case of attrition. Participants will be recruited from the university community and general community of Dunedin, NZ. The study was approved by University of Otago Ethics Committee for Human Participation (REF: 22/049). Informed written consent will be given by each participant prior to their participation.

### 2.2 Study Overview

Each participant will attend four, 30-minutes laboratory sessions, with minimum 1-week apart. The first session will consist of a sucrose detection threshold (DT) task, and a hedonic rating task (used to determine SL and SD according to previous literature). The second and third sessions will be to obtain the *ascending* and *descending* RjTs, the order of which will be counterbalanced across the participants. During the 4^th^ visit, the participants will be given oral instructions for how to complete a 4-day weight Food Record and height and weight will be measured. In addition, the participants will be asked to complete a basic demographic questionnaire and the Dutch Eating Behaviour Questionnaire (DEBQ).

### 2.3 Stimuli

For the sucrose detection threshold task, 10 concentration levels of sucrose solutions will be made by dissolving sucrose (Chelsea, NZ; purity over 99%) in micro carbon-filtered water. The lowest concentration of 0.8g/L is increased by a factor of 1.4 throughout the 10 steps. This concentration range and geometric factor were determined with a pilot study (N=7) prior to the main study. This concentration progression enabled estimation of threshold measures for all of the participants. Water will be used as the blank samples in the detection threshold task.

The hedonic rating task uses five sucrose solutions at the concentration of 30, 60, 120, 240, and 360 g/L. These concentrations had been used in previous literature for classifying SL and SD (Holt, Cobiac, Beaumont-Smith, Easton, & Best, 2000).

For the ascending or descending RjTs tasks, an orange flavoured cordial powder (Baker Halls, NZ) will be used as the base solution. The standard formula with sucrose concentration of 88g/L serves as reference sample. Five ascending and descending levels of sucrose in the orange drink will be made following a geometric series of 1.2. These concentration ranges were tested in the pilot study and were able to produce RjT measures for all participants.

All sucrose solutions will be made in the afternoon before each experiment session, and sub-sampled into brown-coloured glass bottles labelled with 3-digit codes. Sucrose and water samples, 10 mL, will be kept in the refrigerator until 3 hours before the start of the test. At the point of serving, the sample temperature will be approximately 16°C.

### 2.4 Procedures

Participants sucrose detection thresholds will be measured by the method prescribed in ASTM E679. Specifically, each participant is presented with 10 trials of 3-alternative forced-choice task, which comprised one sucrose sample and two blank water samples. The participant is asked to indicate which one is the target (sucrose) sample. Across the trials, the concentration of the sweetened sample is increased according to a geometric progression in an ascending order. A 1-m break is enforced between any two trials.

Hedonic responses for five sucrose aqueous samples will be collected with 15-cm visual analogue scales (VAS). The extreme ends of the scale are labelled as “extremely unpleasant” (0) and “extremely pleasant” (100). The samples will be served to the participant monadically, with an inter-stimulus-interval of 45s. The presentation order of the samples is randomised across the participants following a William’s Latin Square.

Each participant will complete the ascending and descending RjT task across 2 separate sessions. In each RjT session, the participant is presented with a total of 20 trials of 2-alterantive forced-choice (2AFC) preference tests. Each trial comprises one reference sample and one testing sample. The testing samples vary across five different sucrose concentrations. Depending on the task, the sucrose concentration of the testing sample either increases or decreases from the reference sample. Each of the five concentrations will be tested with 4 replicates. With each 2AFC presentation, the participant will be asked to taste the samples monadically and indicate which one they prefer. A response will be enforced when there is no clear preference. All of the above tasks are designed and distributed via *Compusense Cloud* (Compusense, Canada).

For the 4-day weighed Food Record, the participant will be instructed to record all food and beverages consumed over four days, including consecutive weekdays and one weekend day. A Food Record booklet, an electronic scale (M1023, Salter, UK), and a food portion catalogue containing portion sizes (only used when scales cannot be applied, e.g., dining out) will be provided to each participant. In addition, information about alcohol, supplements, or medicines is required, and any event or incidence that may have influenced their eating behaviour is requested to be reported (e.g., attending parties). All participants will be asked to maintain their exercise at a similar level across these testing days. The same researcher will give the instructions to all participants to mitigate inconsistency. This 4-day weighed Food Record booklet has been proofread and approved by a Nutrition academic and New Zealand registered dietitian.

### 2.5 Data analyses

#### 2.5.1 Data processing

Individual detection threshold will be calculated based on ASTM E679. The threshold estimate is the geometric mean across the two concentrations where the participant had their last response reversal from “incorrect” to “correct”. These individual DT measures will be compared to hedonic ratings, ascending and descending RjTs, and included as a baseline measure in a regression model for predicting dietary intake.

Individual hedonic VAS ratings across the five sucrose concentrations will be averaged within participants. Based on methods described in previous research (Holt et al., 2000), the moderate liking value of 50 will be used as a cut-off point for differentiating SL from SD.

Group RjTs will be constructed according to the method in Prescott et al. (2005). For separate SL and SD groups, an aggregated psychometric function based on all responses (all participants and replicates) will be produced by plotting proportion of preference to the reference sample (PC_(Ref)_) against the Log concentration levels. The Log concentration corresponded 75% PC_(Ref)_ will be estimated to be the group RjT.

Similarly, individual RjTs will be estimated with the similar method. Individual psychometric functions will be constructed based on PC_(Ref)_ across the 4 replicates (0%, 25%, 50%, 75%, 100%). The Log concentration at PC_(Ref)_=75% will be extrapolated to be the individual RjT for the testing series. Difference between the ascending and descending Log RjT measure (ΔRjTs) will also be computed for individuals who had reached both measures. This measure represents an acceptable concentration range for the testing group/individual.

Individual 4-day weighed Food Records will be entered into FoodWorks (v2019, Xyris Pty Ltd, Australia), which translates the food records into energy measures (in kJ). The participant’s averaged total energy intake will be used to identify physiologically implausible dietary reports, according to the method prescribed in McCrory, Hajduk, and Roberts (2002). Specifically, individual predicted total energy expenditure (pTEE; based on estimated Basal Metabolic Rates, via Harris–Benedict equation; Roza and Shizgal (1984), and self-reported physical activity level) for the reporting period (see, Lupton et al., 2002 for more information). Using ±2 SD cut-off, individuals whose reported energy intake (rEI) fall outside of 40% - 160% over pTEE are considered as physiologically implausible energy intake and thus discarded. For analyses, individual sucrose intake (kJ and kJ% of total energy intake) will be extracted. Additionally, intake of sweet-taste beverages (g, kJ), defined as non-alcoholic beverage that is sweetened by sugar or sweetener, will also be extracted and examined separately.

#### 2.5.2 Statistical analyses

Univariate analysis of covariates (ANCOVA) will be used to test for differences in DT and hedonic VAS ratings between the SL and SD group, with age, gender, and BMI as covariates. Group RjTs for SL and SD are estimated and contrasted.

A generalised linear mixed-models will be applied to test for differences in individual *ascending* versus *descending* RjTs across the two sweet liker status groups. In this analysis, the *ascending* and *descending* RjTs will be treated as within-subject measures, whereas sweet liker status will be treated as a between-subject measure. A separate ANCOVA will be performed on ΔRjTs to indicate differences between the sweet liker groups. Age, gender, and BMI are also submitted to these models as covariates.

A partial correlation analysis will be applied to assess the relationship between individual ascending and descending RjTs, with age, gender and BMI being controlled for.

Hierarchical regressions will then be performed to determine the role of Ascending and Descending RjTs in predicting sucrose and sweet-taste beverage intake. These models include two steps, with Model I including only participant characteristics (age, gender, BMI), and Modell II include the RjT and ΔRjTs measures.

All analyses will be done with Excel or SPSS. Data of the present study will be deposited in Open Science Framework.

## 3. Discussion

More research focusing on links between individual sweetness perception and sugar intake will be important for elucidating the mechanism underpinning sensory effects on dietary behaviour. Moreover, this line of research can potentially generate relevant information for sugar reduction. While many studies reported hedonic evaluations of specific food products containing full versus reduced sugar contents, there is little systematic understanding of how individuals respond to descending sugar concentrations in real food products.

## Data Availability

All data in the present study will be deposited in the Open Science Framework.

